# Using text mining to track outbreak trends in global surveillance of emerging diseases: ProMED-mail

**DOI:** 10.1101/2020.01.10.20017145

**Authors:** Jingxian You, Paul Expert, Céire Costelloe

## Abstract

**Objectives:** ProMED-mail (Program for Monitoring Emerging Disease, also abbreviated ProMED) is an international disease outbreak monitoring and early warning system. Every year, users contribute thousands of reports that include reference to infectious diseases and toxins, and these reports are then distributed to all subscribers of ProMED. However, the corpus of reports has not been well studied so far. Thus, we propose to apply text mining methods to derive information pertinent to the characterisation of the stage of an epidemic outbreak from the reports.

**Methods:** A retrospective study was conducted in ProMED reports in three steps: reports filtering, keywords extraction from reports and finally word co-occurrence network analysis. The keyword extraction was performed with the TextRank algorithm, keywords co-occurrence networks were then produced using the top keywords from each document and multiple network centrality measures were computed to analyse the co-occurrence networks. We used two major outbreaks in recent years, Ebola 2014 and Zika 2015, as cases to illustrate and validate the process.

**Results:** We found that the information structures extracted at different stages of outbreaks from ProMED are consistent with response strategies as well as situation reports of the World Health Organisation.

**Conclusion:** This study shows that ProMED provides large valuable information to characterise the evolution of epidemic outbreaks. Our research presents a pipeline that can extract and organise this information in a meaningful way. It also highlights the potential for ProMED mail to be utilised in monitoring, evaluating and improving responses to outbreaks.

## INTRODUCTION

ProMED-mail was established and opened to the public in 1994 as an email-based centralising platform to report emerging or re-emerging outbreaks and biologically harmful toxin exposures. Up to now more than 80000 subscribers including, but not limited to, clinicians, media workers and policymakers from at least 200 countries are using it[1]. ProMED now has one main list to post received reports and a number of sub-lists for different languages and areas. Dozens of reports which includes press items, expert opinions, both governmental and non-governmental reports which could be sent by local farmers, clinicians, journalists and researchers reach ProMED’s mailbox every day. In order to ensure the credibility and accuracy of the reports, all the reports are moderated by a group of volunteers before they are posted or sent to subscribers[2]. Thanks to their efforts, ProMED has played a crucial role for several major outbreaks such as SARS in 2003, Ebola in 2014-2016. They are not only the first to report cases in those large-scale outbreaks but also, they record people’s efforts to fight the disease. While more and more experts and specialists have joined the moderators’ team, the moderation is still enormous.

To date, several studies have assessed the efficacy of ProMED as a surveillance system for emerging outbreaks. The first systematic study conducted by Lawrence in 2004 described how ProMED worked and proposed to cooperate with more partners in the future[2]. The authors summarised the achievements that ProMED made from 1994 to 2017[1]. They mentioned that manual moderation of the reports received is both an advantage and a disadvantage since expert opinion might also introduce personal bias. There are also some detailed examinations of ProMED. In 2006, Cowen *et al*. performed a retrospective study of ProMED[3]. They studied the geographical distributions of reports and their outbreak-related timelines finding that a number of diseases were published earlier in ProMED than the largest official outlet of world animal health information, meanwhile, there were also some diseases only reported on ProMED. This study demonstrated the value of ProMED for monitoring diseases. Katherinn *et al*. focused on human melioidosis to evaluate the utility of ProMED for infection control. They observed that ProMED provided daily reports of melioidosis occurrence which helped healthcare workers manage to control the diseases[4]. Besides, Ince *et al*. assessed the reports related to Crimean-Congo haemorrhagic fever virus and validated ProMED as a platform for integrating news of different sources[5]. In summary, they showed that ProMED is holding a prominent position in global health information sharing.

Nonetheless, those previous researches focused on the reports that were posted at the very beginning of an epidemic outbreak, which leaves a large number of reports pertaining to these outbreaks unstudied. To address this situation, we propose using text mining methods to systematically dig into these reports to extract and process all outbreak-related information. We developed a pipeline (Figure 1) to extract information from reports that match given criteria for date range, location range and disease name. To validate the effectiveness of the pipeline, we then conducted two case studies: 2014-2016 Ebola Outbreak in West Africa and Zika Virus Epidemic 2015.

**Figure 1.**
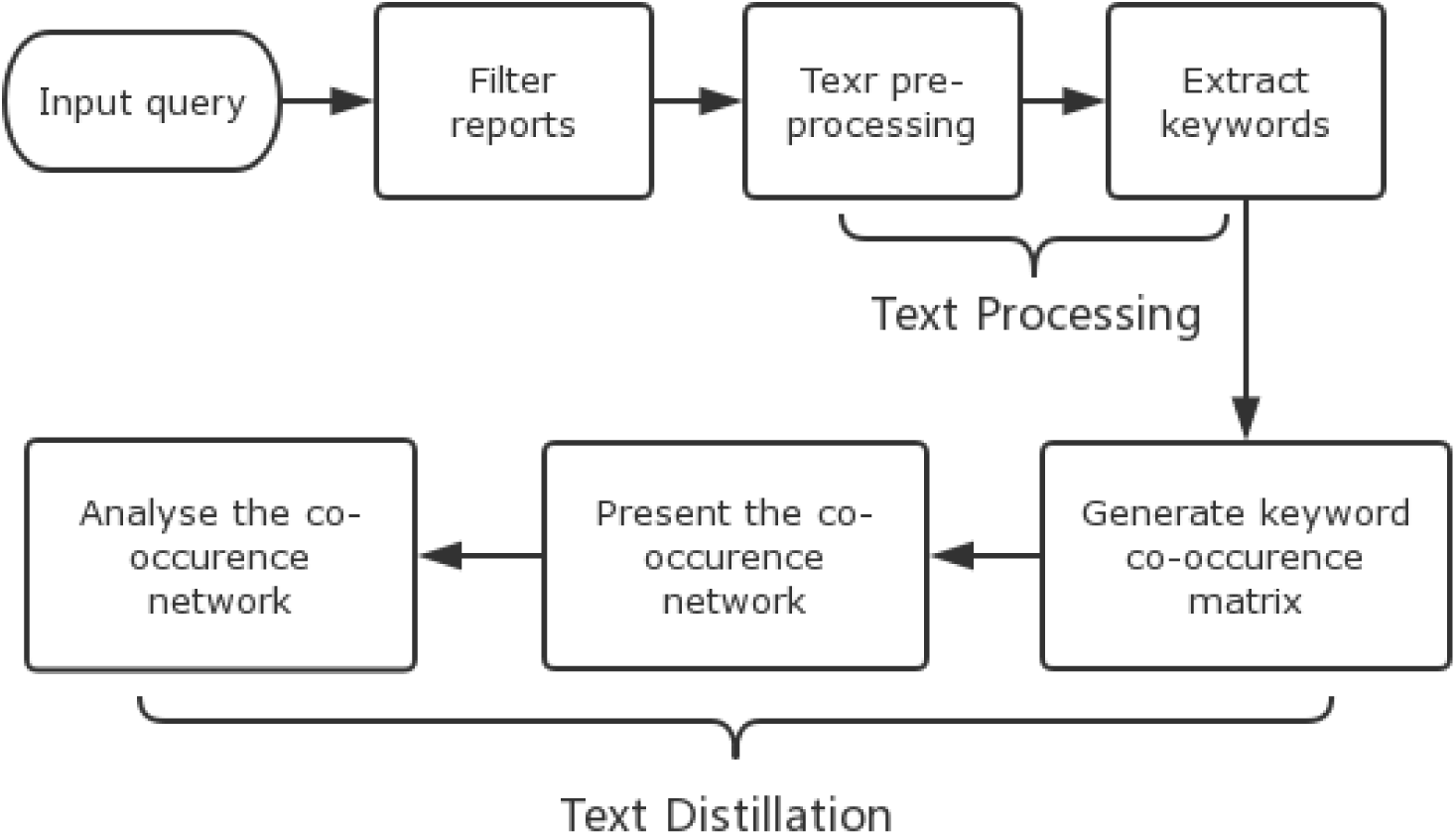
Text mining pipeline including the setting up of criteria and the information extraction from texts to find characterisations of corresponding reports.

An Ebola virus epidemic occurred in Western Africa from 2014 to 2016. ProMED received the first Ebola-related reports from Guinea on March 14, 2014, while it was still suspected of being a case of Lassa Fever. The Ebola virus spread very quickly, and the WHO had to declare a Public Health Emergency of International Concern (PHEIC) to organize and advise governments and individuals to face the challenge. After the outbreak finally ended in June 2016, the WHO summarised the progress of the outbreak and summarised the response in three main phases[6]:

1. August 2014 to December 2014: WHO upgraded the response level at this phase and worked on improving the infrastructure for Ebola treatment.
2. January 2015 to July 2015: Vaccine trials began in Guinea, with an emphasis on how to find new cases and trace contacts more effectively.
3. August 2015 to mid-year 2016: WHO focused on researching the disease itself as well as blocking the transmission of Ebola by improving public health conditions.

Thanks to the timely participation of the WHO and their effective decisions the epidemic was finally brought under control, but tens of thousands of people suffered from physical and mental consequences.

Compared with Ebola, many patients infected with Zika virus do not show particular signs or symptoms, but the virus may bring a series of complications or syndromes such as microcephaly, and other birth defects. The 2015-2016 Zika virus epidemic was well controlled in a relatively short period of time. The PHEIC was declared in February 2016 and ended in November 2016, but local populations still need to be alerted to this disease due to the risk of neurologic complications. We investigated Zika related WHO situation reports and divided this epidemic into three stages for analysis purpose: pre-PHEIC, PHEIC and post-PHEIC.

## METHODS

The pipeline shown in Figure 1 is built in Python. With this pipeline, users can set up criteria by typing countries or continents, diseases, start date and end date to query specific reports. After the reports are filtered, this system will pre-process the relevant reports and extracts keywords, and then perform text distillation by generating co-occurrence networks to refine the knowledge graph as well as analysing the co-occurrences with graph knowledge.

### Text processing – Keyword extraction

The archived ProMED data is saved as a large quote-encapsulated, comma-separated text encoded in MacOS Roman character set. The text pre-processing includes four stages: sentence-splitting; word tokenization; stop words removal and word stemming. We used NLTK (the natural language toolkit package[7], Python) to perform these steps.

The concept of text mining was first systematically summarised by Ah-Hwee Tan in 1999[8]. A framework which included two steps was introduced: text refining and knowledge distillation. The first step, text refining, processes the texts of the corpus under study and extracts features. These will be used at the knowledge distillation stage, where complex relations within a text and among texts forming a corpus can be identified, such as sentiment analysis, fake news identification, etc. Many methods have been developed for text refining, including text summarization[9–11], semantic analysis[12,13], and keyword extraction[14–16], which is the method we use in this study. Current keyword extraction methods can be divided into two categories: machine learning-based and statistics based. Machine learning approaches employ supervised learning methods and thus rely on – potentially only partially - labelled datasets. However, domain-specific labelled datasets are a rare resource, as they require expert field knowledge to yield good performances. On the other hand, statistical approaches, while not as specific, do not suffer from this limitation. Moreover, they do not rely on pre-trained models and are widely used in both academia and industry. There are three common statistical approaches: simple statistics (TF-IDF), topic-based (Topic modelling) and “graph”-based (TextRank).

The ProMED corpus has the following features: 1) High heterogeneity: reports of ProMED can have very diverse resources. 2) imbalanced distribution of topics: ProMED reports always contain disease outbreak information. This implies an uneven distribution in the number of reports for each outbreak, e.g. there are over 4000 reports of Dengue disease, while only hundreds or dozens of alerts for most other diseases. 3) Unlabelled and unstructured: these two features also increase the difficulty of text analysis. The uneven distribution and high heterogeneity lead to the limitation on approaches. Term frequency-based algorithms are not applicable since the frequency of words related to specific diseases may get extremely large during an outbreak and cannot be balanced by inverse document frequency. Taking all these aspects of the corpus into considerations, we chose TextRank as the most suited algorithm for keyword extraction.

TextRank[15] is a graph-based approach which has been shown to work well for many datasets[17,18]. It was derived from the web page ranking algorithm used by Google: PageRank[19]. The starting point is an empty (i.e. zero matrix) adjacency matrix for each word in the text. A sliding window then scans the content of the text: for every pair of words *i* and *j* appearing in the window at the same time, the weight *W*_*ij*_ of the link between words *i* and *j* increased. Once the graph is built, the importance of each word (node) as S(*V*_*i*_) was calculated using the formula:

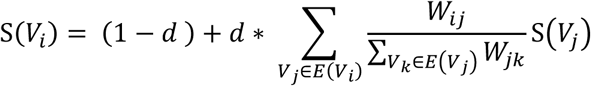

Where d is a damping factor, *E*(*V*_*i*_) is a set of words (nodes) which have edges with *V*_*i*_. The top-k (k = 10) words would be chosen as keywords. The value of d is taken to be 0.85 which has proved effective for keyword extraction[15].

### Text distillation - Keywords co-occurrence network and analysis

After extracting keywords for each text in a corpus, we calculated the co-occurrence matrices of those keywords in all the texts of the corpus. The number of times two keywords are present in a same report is their co-occurrence number and gives a notion of contextual semantic similarity. Co-occurrences matrices can be seen as weighted networks representing the contextual semantic similarities of keywords from a corpus. These networks were imported into Gephi[20] for visualisation and analyses, as shown in Figure 2 and Figure 3. The colour of edges reflects the frequencies of keywords co-occurrence which normalised by the number of reports. The colour bar indicates the range of the normalized frequencies. Since we considered the keyword co-occurrence matrices as networks, we measured different centrality, degree, closeness, eigenvector and betweenness centrality to determine the relative importance of words in the co-occurrence graph. In figures 2 and 3, the sizes of nodes’ labels are proportional to the centrality of nodes measured by degree centrality. We considered the rankings of these words by centrality metrics as the characterisation of the corpus reports under study. A comparison of the rankings of these centrality measures has also been conducted and forms the basis of the discussion.

**Figure 2.**
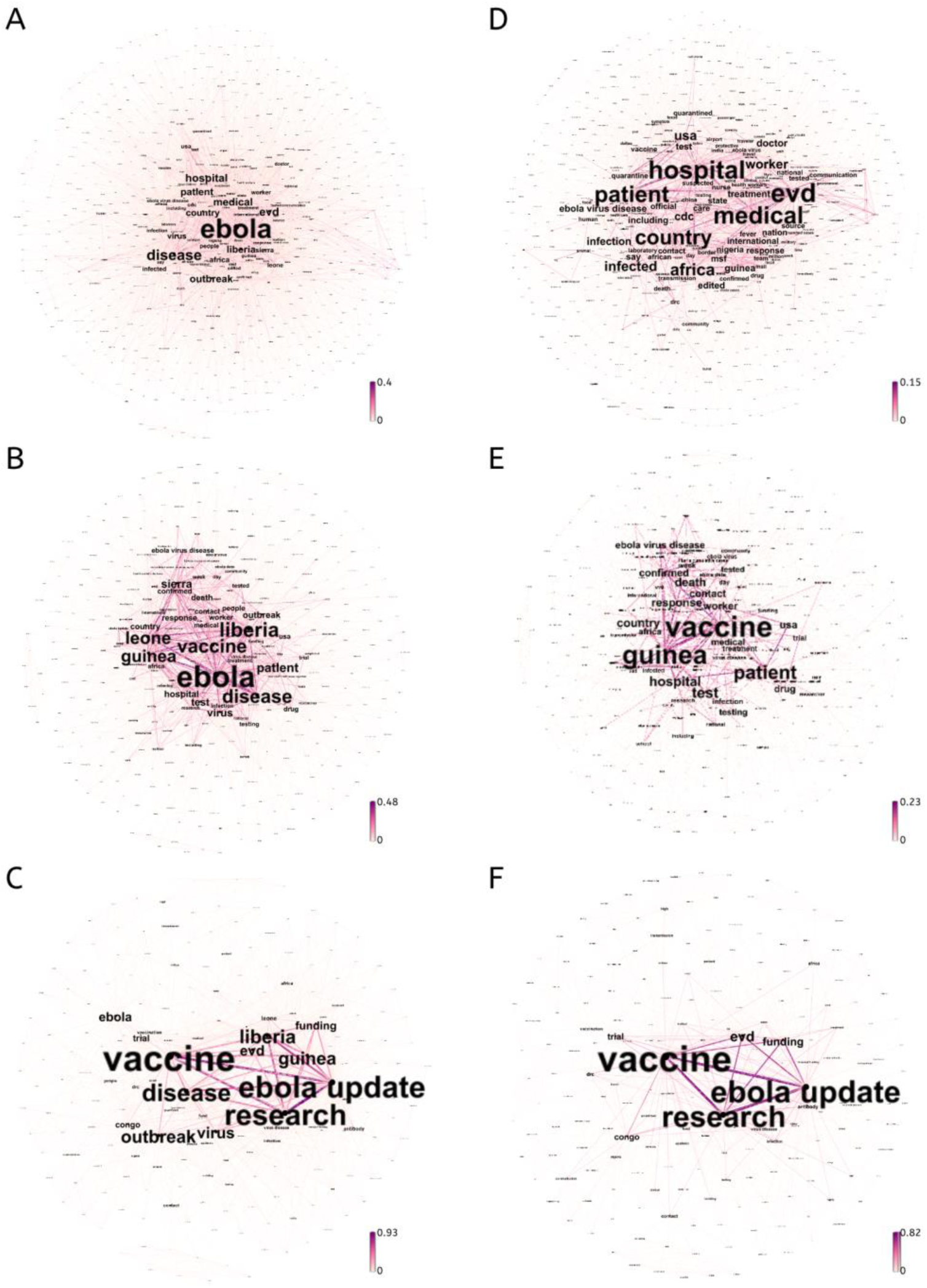
Keyword co-occurrence networks extracted from ProMED reports related to Ebola outbreak 2014. Figure 2.A, Figure 2.B and Figure 2.C are corresponding to PHIEC phase 1,2 and 3, respectively. Figure 2.D, Figure 2.E and Figure 2.F are corresponding to PHEIC phase 1, 2, and 3 after the keyword’s exclusion criteria was applied. The colours of edges reflect the number of co-occurrences between the nodes they are incident to and are normalised by the number of documents in the corpus. The label sizes represent the relative degree centrality of the keywords. The changes of the network are consistent with the WHO’s guidelines. Since the excluded keywords are relatively less important in Figure 2.C, the layouts of Figure 2.C and Figure 2.F are very similar.

**Figure 3.**
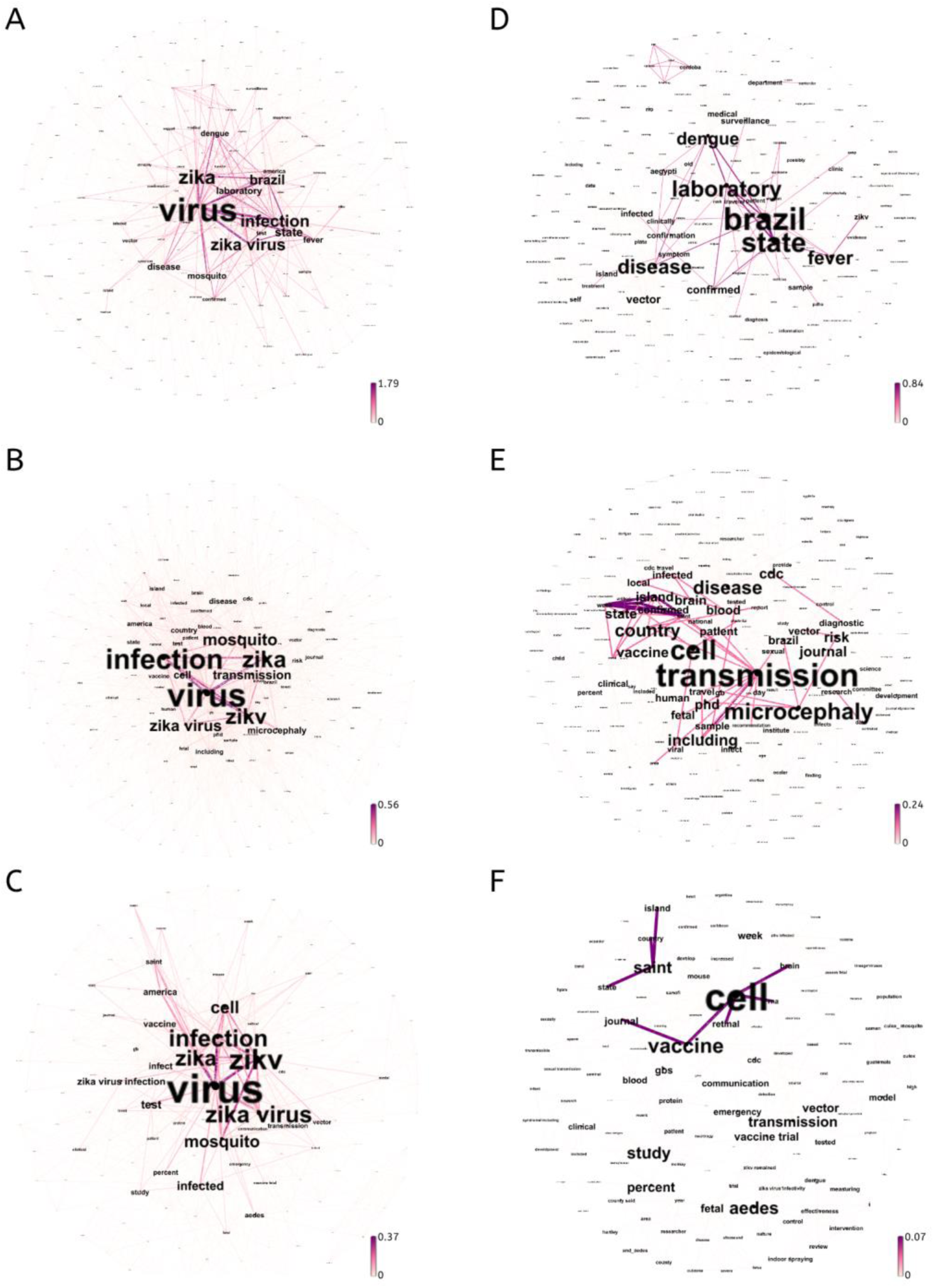
Keyword co-occurrence networks extracted from ProMED reports related to Zika outbreak 2015-2016. Figure 3.A, Figure 3.B and Figure 3.C are corresponding to pre-PHIEC, PHEIC, post-PHEIC stage, respectively. Figure 3.D, Figure 3.E and Figure 3.F are corresponding to these three stages after the keyword’s exclusion criteria was applied. The colours of edges reflect the number of co-occurrences between the nodes they are incident to and are normalised by the number of documents in the corpus. The label sizes represent the relative degree centrality of the keywords. The colours of edges reflect the relative weights and the label size suggests the centrality of the keywords. We can see that the focus of the first stage is the diagnosis of Zika, while the second stage is to block the spread of the disease, and the third stage is the study of the virus.

**Figure 4.**
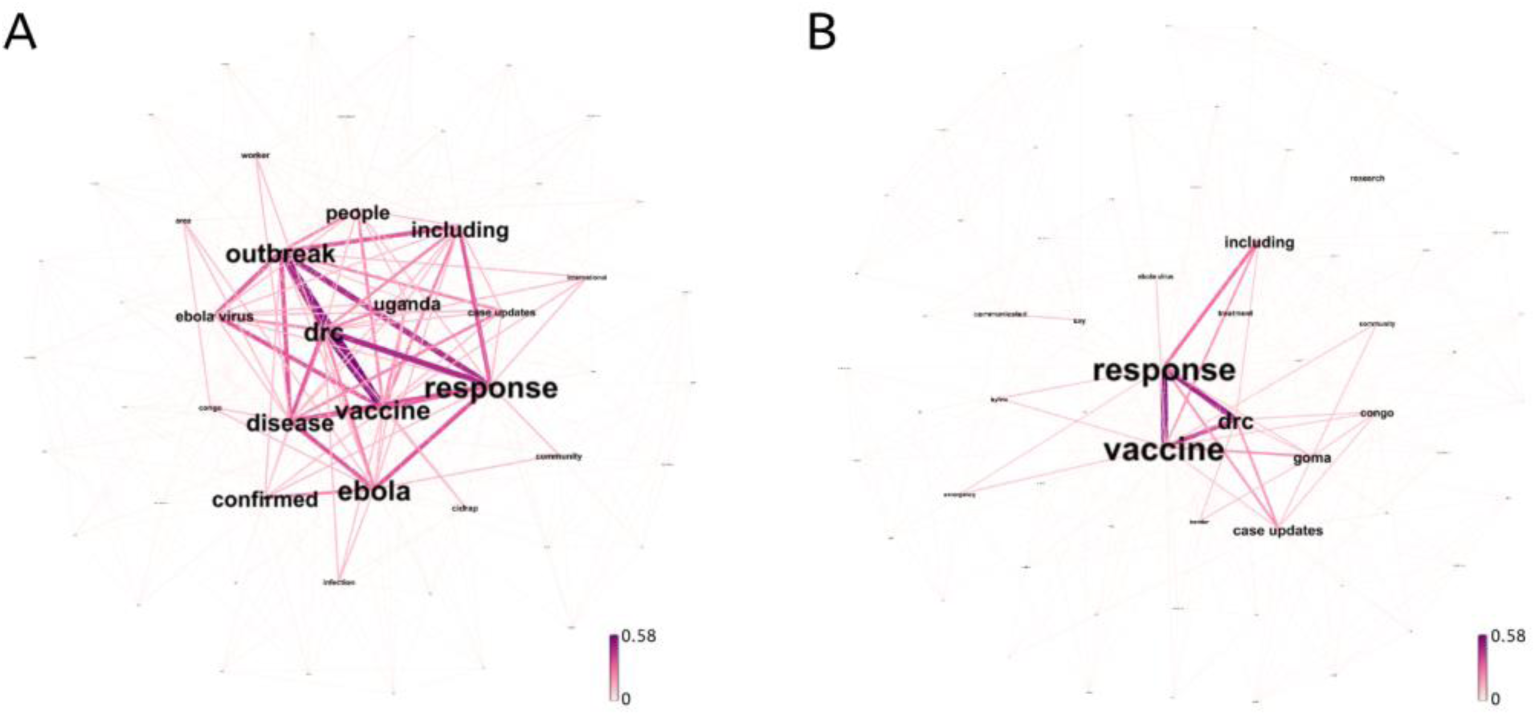
Keyword co-occurrence networks of Ebola-related reports from July 2019 to August 2019. Figure 4.A, original co-occurrence network. Figure 4.B co-occurrence network after “Ebola”, “outbreak”, “disease”, “virus” and “people” removed. The colours of edges reflect the number of co-occurrences between the nodes they are incident to and are normalised by the number of documents in the corpus. The label sizes represent the relative degree centrality of the keywords. It shows that the two priorities in the current epidemic in July and August 2019 are the response to the outbreak and the study of vaccines.

## RESULT

### Text processing - Keyword extraction from single reports

We extracted keywords and kept the 10 keywords with the highest TextRank scores of each report. Table 1 shows the keywords extracted from 3 randomly chosen reports. Alert 4735420 is about Pertussis cases confirmed in northern Wisconsin. The algorithm extracted keywords including location, disease name and other critical information like vaccine, which arguably concisely summarises the most important aspects of the report. Likewise, vector, disease and location were also successfully extracted from alert 4736083, which was referring to a confirmed hantavirus case in Maule Region. However, in terms of the alert 4735363, which is reporting a Lassa fever outbreak that happened in Ogun State, the algorithm failed to extract complete location name but only extract ‘state’ instead.

**Table 1:** Extracted keywords from the 3 isolated ProMED reports 4735420, 4736083 and 4735363.

| Alert 4735363 | Alert 4735420 | Alert 4736083 |
| --- | --- | --- |
| Rodent | Wisconsin | Rodent |
| Mestomes | Immune | Hantavirus |
| Lassa Fever | Report | Positive |
| Confirmation | Country | Previous |
| State | Pertussis | Treatment measure |
| Nosocomial | Cough | Andes |
| Threat | School | Maule |
| Project | Diphtheria | Mod |
| Epicore | Year | Contact |
| Surveillance | Vaccine | physical |

### Text distillation – Text mining from a corpus of reports

#### Case study 1: Ebola virus disease

We grouped and analysed the reports for each period defined by the WHO and compared the most central keywords of each stage, thus three subcorpora have been analysed. The co-occurrence networks can set out in Figure 2. Table 2 shows the top 10 words with the highest degree centrality in the co-occurrence networks representing each time period. Words which ranked top 10 in all three networks were excluded as they are characterising the disease in general and not the stage it is at. For example, ‘Ebola’ ranked 1^st^, 1^st^,10^th^ in three stages, respectively, so we removed it from all the three stages. The words that are excluded are “Ebola”, “disease”, “outbreak”, “Liberia”, “Sierra”, “Leone”, “virus” and “people”. We then excluded the nodes representing these words from the original networks, and the results can be compared in Figure 2. We also calculated other centrality measures (Supplementary Table 5-7) and the top 10 words in the ranking results were identical for each after the exclusion was applied.

**Table 2:**
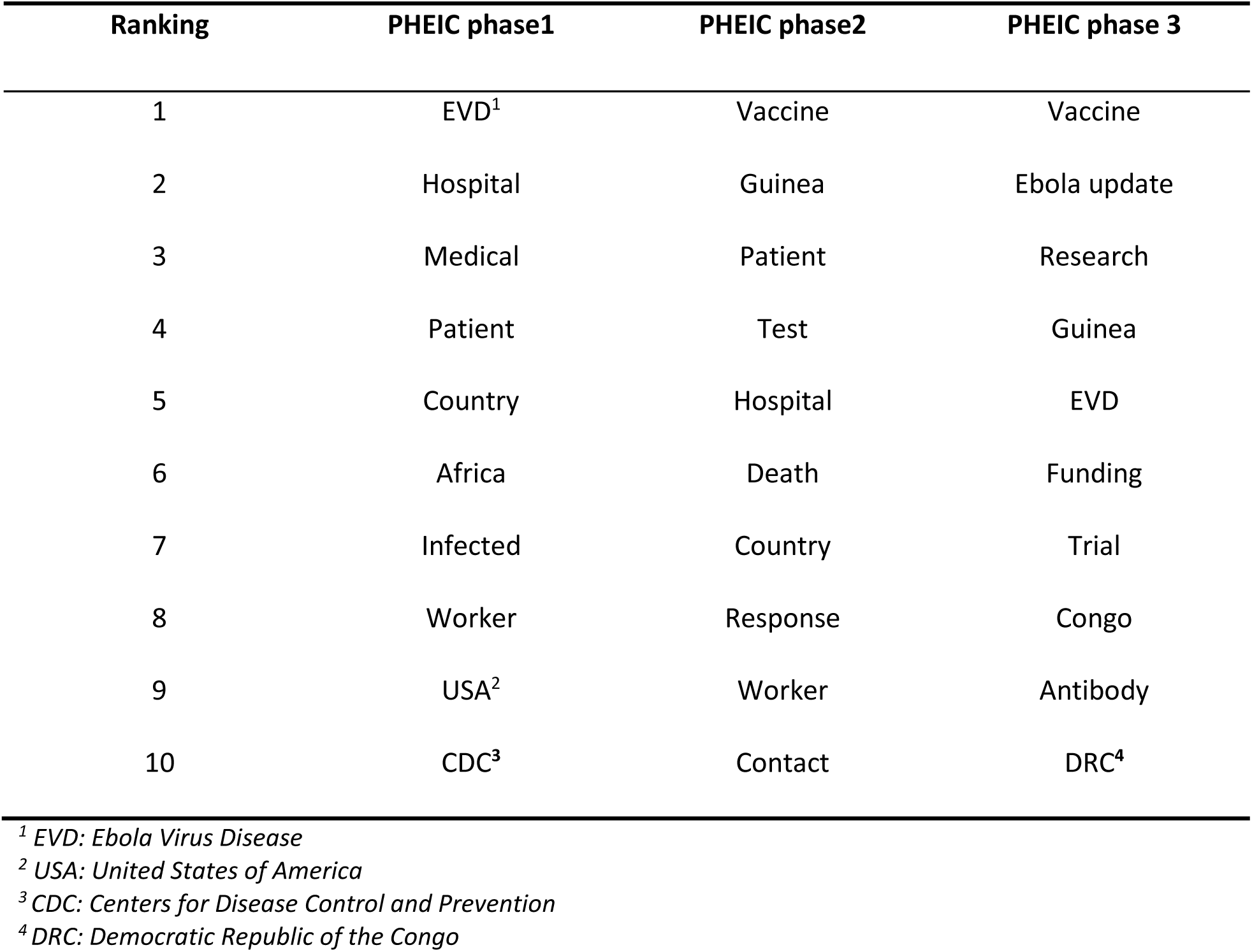
Extracted high degree centrality keywords from 3 PHEIC phases of 2014-2016 West Africa Ebola Outbreak

#### Case study 2: Zika virus disease

As it broke out in South America, many ProMED reports were in Portuguese or Spanish - among all the reports which mentioned “Zika” in the subject line in 2016, 162 of them were in Portuguese, and 140 were in Spanish compared to only 65 were in English. Although our pipeline does not have a limitation of languages, since we are not familiar with Portuguese or Spanish, and in order to avoid potential translation problems, we only analysed the report in English.

The keywords co-occurrence networks corresponding to the stages are shown in Figure 3. We applied the same removal criterion with the Ebola study to refine the ranking of the keywords which is shown in Table 3. The removed words are “Zika”, “virus”, “infection”, “Zika virus”, “mosquito”, “America”, “test”.

**Table 3:** Extracted high degree centrality keywords from pre-PHIC stage, PHEIC stage and post-PHEIC stage of 2015 Zika Outbreak

| Ranking | Pre-PHEIC | PHEIC | Post-PHEIC |
| --- | --- | --- | --- |
| 1 | Brazil | Transmission | Cell |
| 2 | State | Cell | Vaccine |
| 3 | Laboratory | Microcephaly | Saint |
| 4 | Disease | Disease | Study |
| 5 | Fever | Country | Percent |
| 6 | Dengue | Including | Aedes |
| 7 | Vector | Risk | Transmission |
| 8 | Confirmed | Vaccine | Vector |
| 9 | Surveillance | CDC | GBS<sup>5</sup> |
| 10 | Sample | State | Vaccine trial |

<sup>5</sup> GBS: Guillain-Barré syndrome

## DISCUSSION

As expected, the co-occurrence networks formed by the keywords extracted by the TextRank algorithm is an excellent characterisation of the trends. Specifically, we obtained summary words for each phase based on centrality ranking and they matched the facts of the same period of time which were independently defined by other organizations. In our first case study, “hospital” and “worker” rank 2^nd^ and 8^th^ in the first phase which is corresponding to the WHO’s strategy – to improve local infrastructure. This correspondence can also be observed in the two other phases: “hospital” and “worker” rank lower and “vaccine” becomes predominant, words “response” and “contact” appear in the second phase while “funding”, “research”, “trial” and “antibody” represent the third phase. This correlation also suggests that people do follow the WHO’s strategy to confront the epidemic. Similarly, ProMED reports and the development of the Zika epidemic are also in good correspondence. First, words “Dengue”, “Vector” and “Laboratory” can be seen at Pre-PHEIC stage. We then further investigated relevant reports and found that this was because dengue and Zika virus disease often had the same vector and similar symptoms; thus, hospitals were in need of laboratory testing to determine the disease. Second, from the Table 3, we can see “Microcephaly” in PHEIC stage, as ProMED reports of this stage suggested the correlations between the Microcephaly and Zika virus disease. And this correlation later became a scientific consensus. Third, “Vaccine” first appears in PHEIC stage at the 8th place and becomes predominant – 2nd place in the Post-PHEIC stage along with “study” – 3rd place.

### Natural language processing (semantic level)

The current pipeline may be somewhat limited by the quality of extracted keywords. For example, it failed to detect synonyms like “evd” and “ebola virus disease”. It also failed to extract phrases like “Ogun state”. Since the ultimate goal of ProMED is to provide an early warning system, if we stick to the automated process, an extremely high accuracy approach is needed. As we mentioned above, TextRank is currently the most suitable unsupervised algorithm for current ProMED dataset. In order to improve reliability, two methods that could be used: first, to employ a pre-trained word embedding model for NER task (Named Entity Recognition). The difficulty lies in the lack of the model. Few models were trained on corpus similar to epidemical reports. This gives rise to a second obstacle – the heterogeneity of ProMED reports may affect the density of entities which means it may also affect the accuracy. The second option is to use supervised algorithms. In this case, it would be necessary to heavily invest in labelling the data accurately. Incorporating these approaches in future work would facilitate the development of an automated system to sort the reports and to extract critical information, accurate to information sources, species, location, lethality and even disease vector.

Another option is to rely on human summary of the reports, bypassing the need for keyword extraction. ProMED may request that the submitted report must indicate the information we need in a certain format, leaving only the distillation stage. Indeed, the reports will now be edited by the moderators to add the subject line containing some information like location name, disease name before they are posted. However, the original reports may still be ambiguous in some vital aspects. Thus, it would be necessary for ProMED to provide guidelines for summarizing documents. This could alleviate moderators’ pressure to some extent and reduce the risk of human bias by reviewing the reports at both the submission and moderation levels.

### Epidemiology analysis

A transport data-based, network-driven epidemiology prediction model was developed by Colizza *et al*. in 2006[21]. This system relies on the global airline data and local commuting data of more than 40 countries to calculate the probability of infection and has been successfully applied to several outbreaks including 2014 Ebola outbreak[22], 2014 MERS outbreak[23] and 2015-2016 Zika outbreak[24]. Combining this model with information extracted from ProMED reports could lead to the development of a real-time epidemic alert system. Based on immediate reports, such a system should give an accurate estimate of the outbreak probability of potential infectious diseases. Local commuting data which was used in the previous work can be replaced by the travel time data from the Malaria Atlas Project (MAP) which would be more suitable for epidemic modelling[25]. With MAP dataset, we can estimate travel times between locations, which could be a more comprehensive and accurate global measurement than commuting data.

### Ebola 2019 outbreak

Democratic Republic of the Congo declared a new Ebola outbreak on August 1^st^, 2018 and a PHEIC was declared on July 19^th^, 2019. We then examined the reports from July 2019 to August 2019 and we removed the same keywords that were excluded in the reports related to the previous epidemics. The result was compared with the 2014 Ebola outbreak by Jaccard similarity coefficient. Table 4 presents the summary Jaccard coefficients. As can be seen in the table, the current epidemic in Congo has a closer correlation with the final PHEIC stage of the 2014 Ebola outbreak compared with two other stages. This suggests that lessons have been learned from the experiences of earlier epidemics: the response was faster and the epidemic better controlled.

**Table 4:**
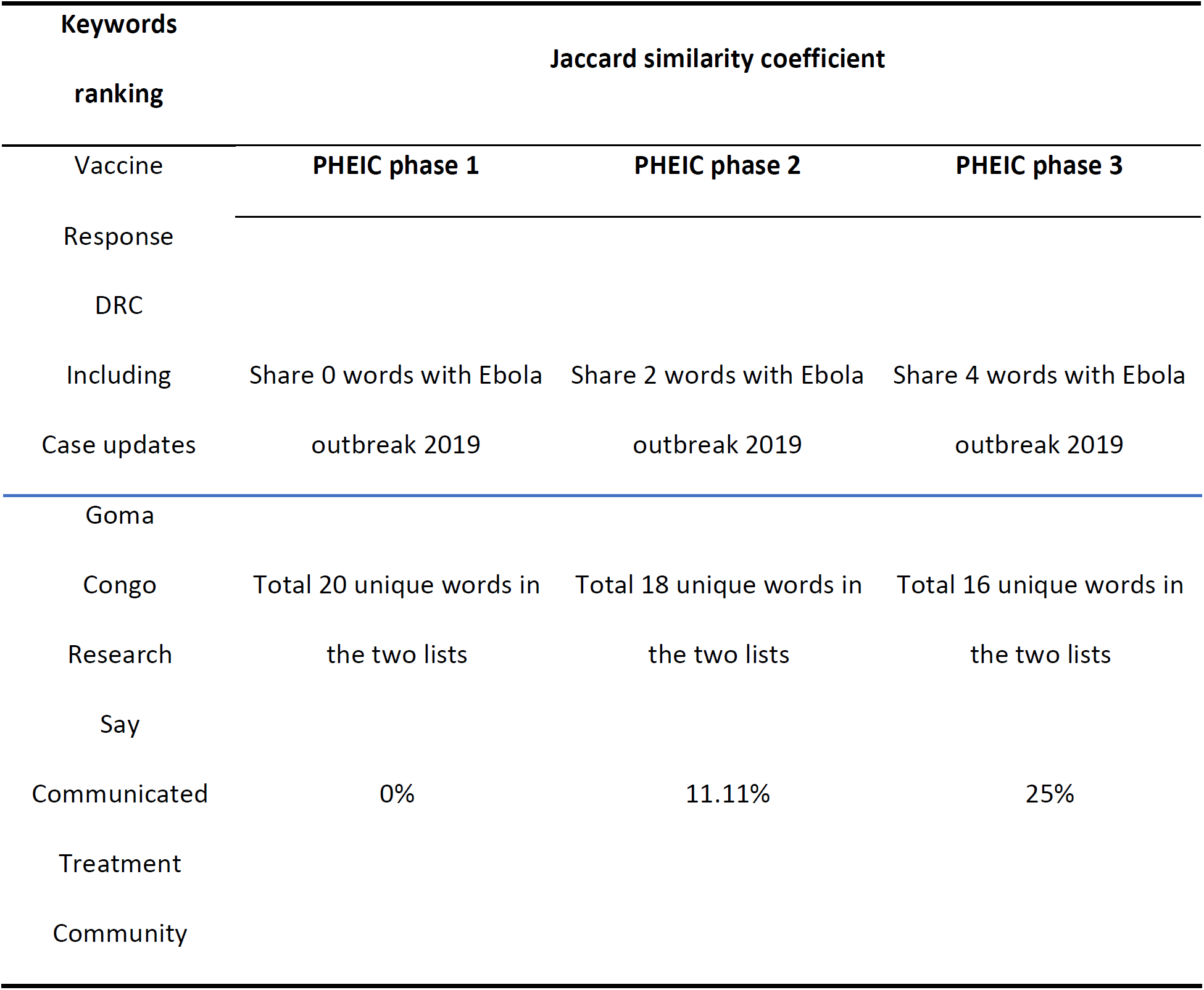
Extracted high degree centrality keywords from 2019 Ebola Outbreak

| Keywords<br>ranking | Jaccard similarity coefficient |  |  |
| --- | --- | --- | --- |
|  | PHEIC phase 1 | PHEIC phase 2 | PHEIC phase 3 |
| Vaccine |  |  |  |
| Response |  |  |  |
| DRC |  |  |  |
| Including | Share 0 words with Ebola | Share 2 words with Ebola | Share 4 words with Ebola |
| Case updates | outbreak 2019 | outbreak 2019 | outbreak 2019 |
| Goma |  |  |  |
| Congo | Total 20 unique words in | Total 18 unique words in | Total 16 unique words in |
| Research | the two lists | the two lists | the two lists |
| Say |  |  |  |
| Communicated | 0% | 11.11% | 25% |
| Treatment |  |  |  |
| Community |  |  |  |

## CONCLUSION

In this study, we implemented an unsupervised algorithm in a large corpus of epidemiological reports and further investigated the knowledge structure using keyword co-occurrence network. Through two case studies, we showed that our pipeline can be used for analysis of epidemic response trends and potentially as an early stage alerting system. Since the trends are consistent with the WHO’s strategies, we also conclude that ProMED, as an independent NGO, can help governments and institutions better understand the epidemic situation, formulate policies and follow their implementation.

## Data Availability

All data is available to public online.

https://promedmail.org/

## ACKNOWLEDGEMENT

The authors would like to acknowledge and thank Dr David Birch for valuable advice in both text mining and network visualization.

## COMPETING INTERESTS

No competing interest.

## CONTRIBUTION

CC proposed the study, contributed to data acquisition as well as experiment design, and provided useful comments to the manuscript. JY conducted the majority of the experiments and drafted the manuscript. PE provided critical suggestions on data analysis and edited the manuscript detailly.

## FUNDING

This research was funded by the NIHR Imperial Biomedical Research Centre (BRC). The views expressed are those of the author(s) and not necessarily those of the NIHR or the Department of Health and Social Care.

Jingxian You and Paul Expert were supported by the NIHR Imperial Biomedical Research Centre (BRC). Céire Costelloe holds a personal NIHR Career Development Fellowship (NIHR-CDF-2016-50)

## Notes

### Competing Interest Statement

The authors have declared no competing interest.

